# Quantification of Optical Coherence Tomography Angiography in Age and Age-related Macular Degeneration using Vessel Density analysis

**DOI:** 10.1101/19003053

**Authors:** Ehsan Vaghefi, Sophie Hill, Hannah M Kersten, David Squirrell

## Abstract

**Purpose:** To determine whether vessel density (VD) as measured by optical coherence tomography angiography (OCT-A) provide insights into retinal and choriocapillaris vascular changes with ageing and intermediate dry age related macular degeneration (AMD).

**Methods:** Seventy-five participants were recruited into three cohorts; young healthy (YH) group, old healthy (OH) and those at high-risk for exudative AMD. Raw OCT and OCT-A data from TOPCON DRI OCT Triton were exported using Topcon IMAGENET 6.0 software, and 3D datasets were analysed to determine retinal thickness and vessel density.

**Results:** Central macular thickness measurements revealed a trend of overall retinal thinning with increasing age. VD through the full thickness of the retina was highest in ETDRS sector 4 (the inferior macula) in all the cohorts. Mean VD was significantly higher in the deep capillary plexus than the superficial capillary plexus in all ETDRS sectors in all cohorts but there was no significant difference noted between groups. Choriocapillaris VD was significantly lower in all ETDRS sectors in the in the AMD group compared with the YH and the OH groups.

**Conclusions:** Retinal vessel density maps, derived from the retinal plexi are not reliable biomarkers for assessing the ageing macular. Our non-proprietary analysis of the vascular density of the choriocapillaris revealed a significant drop off of VD with age and disease but further work is required to corroborate this finding. If repeatable, choriocapillaris VD may provide a non-invasive biomarker of healthy ageing and disease.

**Brief Summary:** In this manuscript, we have studied the potential of retinal vessel density as measured by optical coherence tomography angiography (OCT-A), as a biomarker for detection of high-risk of developing exudative age-related macular degeneration (AMD).

## Introduction

Currently we lack a reliable, non-invasive vascular biomarker to monitor both the “healthy” ageing and disease progression of age related macular degeneration (AMD). Optical coherence tomography angiography (OCT-A) represents a novel, non-invasive, dye-less retinal vascular imaging technique that can be rapidly acquired during a clinical consultation. OCT-A is a progression of OCT, using repeated B-scans to detect motion contrast^1^. Based on the assumption that this represents the movement of erythrocytes travelling through the retinal blood vessels, the retinal vasculature can therefore be visualised.^2^ Previous studies have shown the value of this imaging technique in the evaluation of glaucoma, diabetic retinopathy (DR), retinal vein occlusions and AMD^3^.

The laminar structure of the retina lends itself to segmentation analysis. Histologically, the retina has four retinal capillary plexuses, within the macular only three of these are considered; the superficial, deep and intermediate capillary plexuses^4, 5^. When the macular vasculature is imaged using OCT-A the vascular layers are segmented automatically by the software. This results in the merging of the intermediate with the deep capillary plexus^3^. The regions of interest identified on the OCT-A therefore include the superficial capillary plexus (SCP) and the deep capillary plexus (DCP)^3^. The SCP consists of the vasculature within the retinal nerve fibre layer (RNFL) and ganglion cell layer (GCL). The DCP represents the vascular plexuses present at two locations: These plexuses are at the border of the inner nuclear layer and the outer plexiform layer, and the border of the inner plexiform layer and inner nuclear layer^6^.

OCT-A images can be subjectively appraised for the presence of disease or post processed to produce quantitative data that can be evaluated objectively. A number of studies^7, 8^ have used perfusion indices such as vessel density and flow index^9^ as a method of quantitatively analysing OCT-A images. Vessel density is defined as the “percentage area occupied by vessels in the segmented area”^9^.Flow index is defined as “the average decorrelation values in the segmented area”^9^. It has been proposed that these indices could be of value in monitoring disease progression in AMD and DR^7, 10, 11^. When measuring vessel density, both vessel length and diameter need to be considered. Poor perfusion can also result in vessel dilation, therefore vessel density may not give complete information about the retinal vascular status^12^.

There are a wide range of OCT-A devices available, each using a variety of algorithms to capture retinal images. These include OCT-based optical micro-angiography (OMAG); split-spectrum amplitude decorrelation angiography (SSADA); OCT angiography ratio analysis (OCT-ARA); speckle variance; phase variance; and correlation mapping^13^. Specific OCT-A models such as the AngioVue (Optovue, Inc., Fremont, CA, USA) have inbuilt software for perfusion indices calculation^14^. However, the majority of studies have applied post image analysis in a variety of methods after the images are exported from the OCT-A instrument^2^. This heterogeneity in the image capture and analysis means that data calculated using different algorithms are not directly comparable, due to systemic difference and poor agreement^2^. Therefore, external normative databases are unreliable and longitudinal monitoring of disease using vessel density requires scans to be performed using the same instrument, scan location and algorithm^2^.

The aims of this study are to compare the macular vessel density values for; young healthy, old healthy, and patients with intermediate dry age related macular degeneration at high risk of progression to exudative AMD, and to determine if these perfusion indices could usefully serve as a biomarker for both “healthy” ageing and the identification of disease risk.

## Materials and Methods

Seventy-five participants were recruited through Auckland Optometry Clinic and Milford Eye Clinic, Auckland, New Zealand. All participants provided written informed consent prior to imaging. Ethics approval (#018241) from the University of Auckland Human Participants Ethics Committee was obtained for this study. This research adhered to the tenets of the Declaration of Helsinki.

Participants were divided into three groups, young healthy (YH) group, old healthy (OH) group and the high-risk intermediate AMD group. Twenty participants were recruited into the YH group, twenty-one were recruited into the OH group, and thirty-four recruited into the AMD group. A comprehensive eye exam was conducted on each participant prior to the OCT and OCT-A scans in order to determine high contrast BCVA and the ocular health status of the fundus. Patients with any posterior eye disease that could potentially affect the choroidal or retinal vasculature including, but not limited to glaucoma, polypoidal choroidal vasculopathy, DR, hypertensive retinopathy, and high myopia (≥6D), were excluded from the study. Patients with any history of neurological disorders were also excluded. The Young Healthy group consisted entirely of individuals between the ages of 20 and 26 and a best corrected visual acuity of ≥6/9 in the eye under test. The “Old Healthy” group consisted of individuals over the age of 55 years and who had and a best corrected visual acuity of ≥6/9 in the eye under test. The “AMD group” consisted entirely of patients with high risk intermediate dry AMD. This being diagnosed if the individual had at least two of the following risk factors; reticular pseudodrusen, established neovascular AMD in the fellow eye, confluent soft drusen with accompanying changes within the retinal pigment epithelium. In order to ensure that all patients in the “AMD cohort” could maintain fixation during OCT-A imaging, only those patients with a best-corrected visual acuity (BCVA) of 6/15 or better were enrolled. The mean age of the participants in the YH group, OH and AMD participants were 23 ± 3, 65 ± 10 and 75 ± 8 years respectively. Only one eye of each patient was used for the analysis, and if the patient had both eyes scanned, the OCT-A scan that was the better quality (assessed subjectively by the clinical grader) of the two was used. Mean best-corrected visual acuity for the YH, OH and AMD group were 6/6, 6/9, and 6/12 respectively.

The ocular health of all participants was assessed at Auckland Optometry Clinic, by a registered optometrist, prior to enrolment in the study. The macular status of patients enrolled into the AMD group was assessed separately by an experienced retinal specialist (DS).

### Data collection protocol

Participants were dilated with 1.0% tropicamide if the pupils were deemed to be too small for adequate OCT scans. Intraocular pressures were measured before and after dilation by iCare tonometry. OCT/OCT-A scans, fundus photography and clinical measurements were all performed at the University of Auckland Optometry Clinic, in a single session.

### SS-OCT-A device and scanning protocol

The swept source OCT-A device (Topcon DRI OCT Triton, Topcon Corporation, Japan) was used to obtain the OCT and OCT-A scans. A macular line and 3×3mm^2^ OCT, and 3×3mm^2^ OCT-A scan were performed on each participant. All scans were cantered on the fovea, and retinal layers were identified using the IMAGENET 6.0 automated layer detection tool so that the superficial (SCP) and deep (DCP) capillary plexuses could be evaluated. The quality of the generated layers were checked manually. Scan quality was evaluated at the time of acquisition, and repeated if required.

### Quantitative analysis of OCT-A Images

Raw OCT and OCT-A data were exported using Topcon IMAGEnet 6.0 software. Since OCT-A images are only qualitative, we chose to normalize the OCT-A datasets, prior to further analysis. These datasets were then correlated with vessel density in each retinal sector. OCT-A 3D datasets were analysed in three different ways (outlined below), to investigate the retinal and choroidal thickness and vessel density (VD).

#### 1. ETDRS-specific

The nine Early Treatment Diabetic Retinopathy Study (ETDRS) sectors were computationally generated and centred manually by the clinical examiner, on the fovea (using enface OCT-A) for each participant. In some instances, the macular region was not in the centre of the enface 3×3mm^2^ OCT-A scan. Hence, after manual adjustment of the ETDRS central positioning on the fovea, the outer ETDRS regions (i.e. 6-9) could have been positioned outside of the imaging area. Therefore, in this study in which were interested in assessing the role vessel density may play as a potential biomarker for ageing and disease of the central macular, we only used ETDRS regions 1 to 5 for consistency in our analysis. In this process, the OCT-A signal is extracted and saved for each ETDRS region, superimposed on every OCT-A enface layer. In other words, all the 3×3mm^2^ OCT-A layers through the retinal thickness are combined to create an OCT-A volume dataset, 920 of them per 3×3mm^2^ scan, using the Topcon DRI OCT. Finally, the ETDRS regions are manually applied to each of the 920 enface OCT-A images and OCT-A normalized data per region are extracted (see Figure 1). This method has been recently published^15^.The averaged normalized vessel density (VD) of the full thickness retina and choriocapillaris of each ETDRS region was then extracted from every participant for further analysis. Normalisation was performed by converting the raw data to percentage of VD in all ETDRS regions, throughout the thickness of the retina.

**Figure 1:**
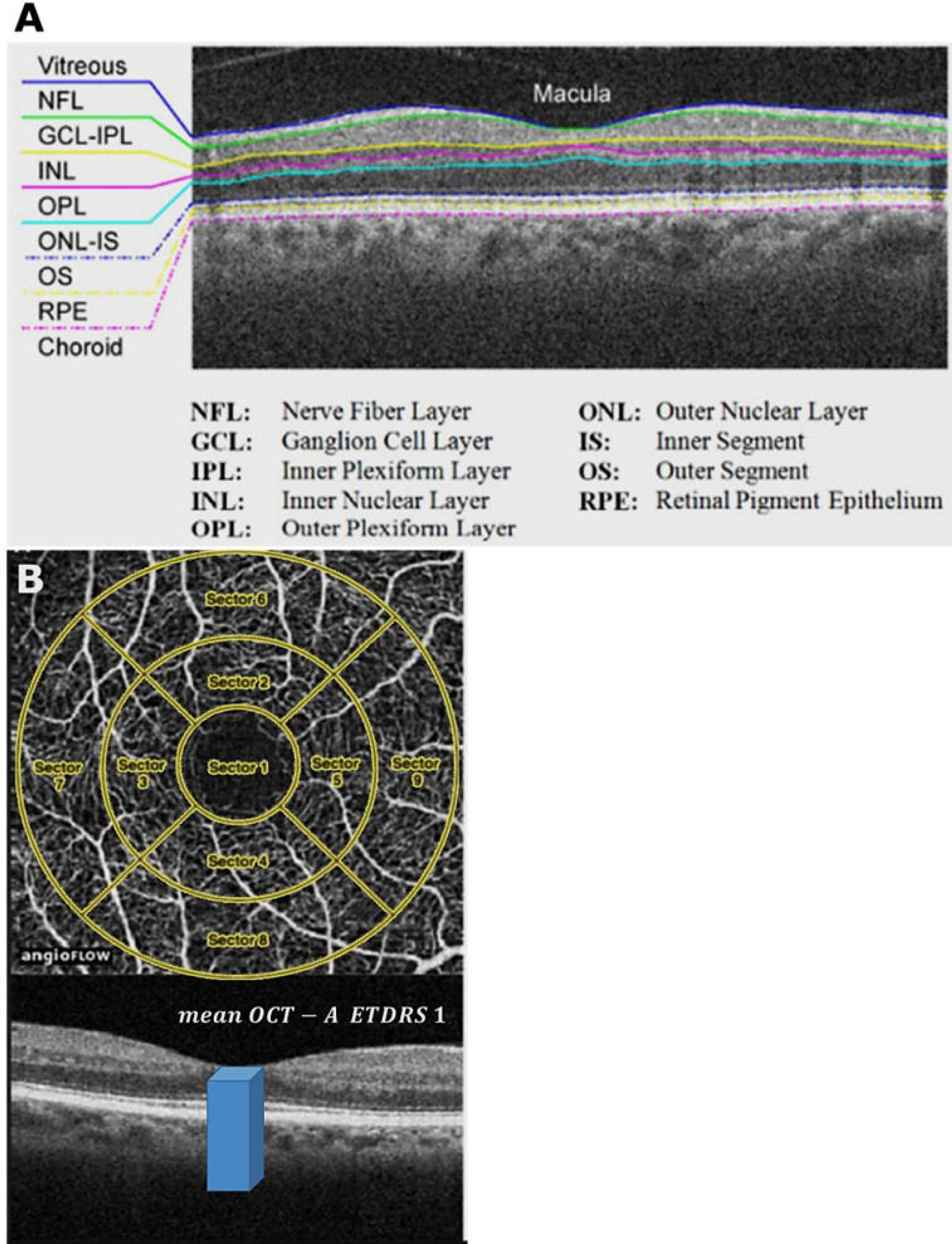
Retinal layers were automatically identified by IMAGENET 6 software (A) the SCP and DCP were then isolated from layers 1 and 2, and 3 and 4 respectively. ETDRS regions 1-9 are superimposed onto the enface OCT-A images and manually adjusted so that sector 1 was centred on the fovea. In this study we selected sectors 1-5 (B).

#### 2. SCP, DCP and Choriocapillaris

Using the same ETDRS regions as above, the VD was extracted from layer 1 and 2 for the SCP calculation, layers 3 and 4 for the DCP and layer 8 for the choriocapillaris. Averaged normalized VD from the SCP, DCP and choriocapillaris was then extracted for each participant, in the three groups.

#### 3. Retinal and choroidal thickness

In addition to OCT-A data, we also recorded the foveal thickness and subfoveal choroidal thickness from the OCT B-scans.

### Statistical analysis

MATLAB programming software and custom-written code was used to import and analyse the data, as detailed above. Statistical analysis was performed using MATLAB Statistics Toolbox. Due to unequal sample sizes between the sample groups, we performed the two sample non-parametric Kolmogorov and Smirnov test, which is one of the most general nonparametric methods for comparing two samples. The Kolmogorov and Smirnov method assumes that the data in both groups follow Gaussian distributions. Significant differences between groups were defined as D<0.565 for the Kolmogorov and Smirnov test.

## Results

### Mean age, central macular thickness, and foveal avascular zone diameter

Seventy-five participants were included in this study of which 20 were in the Young Healthy group (23 ± 3 years old), 21 in the Old Healthy group, (65 ± 10 years old) and 34 in the AMD group (75 ± 8 years old). OCT data was segmented by IMAGEnet 6.0 software, and the mean central macular thickness measurements for the three participating cohorts was extracted: YH 217.4 ±9.28 μm, OH 209.8 ±6.13 μm, AMD 174.2 ±20.3 μm. Horizontal diameter of the foveal avascular zone (FAZ) was also measured from each OCT and mean FAZ measurements were calculated: YH 672.5 ± 73.4 μm, OH 577.7 ± 98.8 μm, AMD 639 ± 126.2 μm.

### Vessel density measurements: Full retinal thickness

Table 1 shows the mean VD throughout the ***full retinal thickness***, for each ETDRS sector. In all groups the highest VD was found in sector 4, the inferior macula. Lowest vessel density in all groups was in sector 1 the foveal/parafoveal region. A statistical difference between the vessel density in Sector 1 was found between all the groups, with the vessel density being higher in the AMD cohort (29.6±19.8) compared to the Young Healthy (7.5±6.6) and Old healthy groups (14.7±9.68). Analysis of the vessel density of ETDRS sectors 2-5, revealed that there was a statistical difference in the VD between the AMD group and both the YH and OH groups; but not between the YH and OH groups.

**Table 1:**
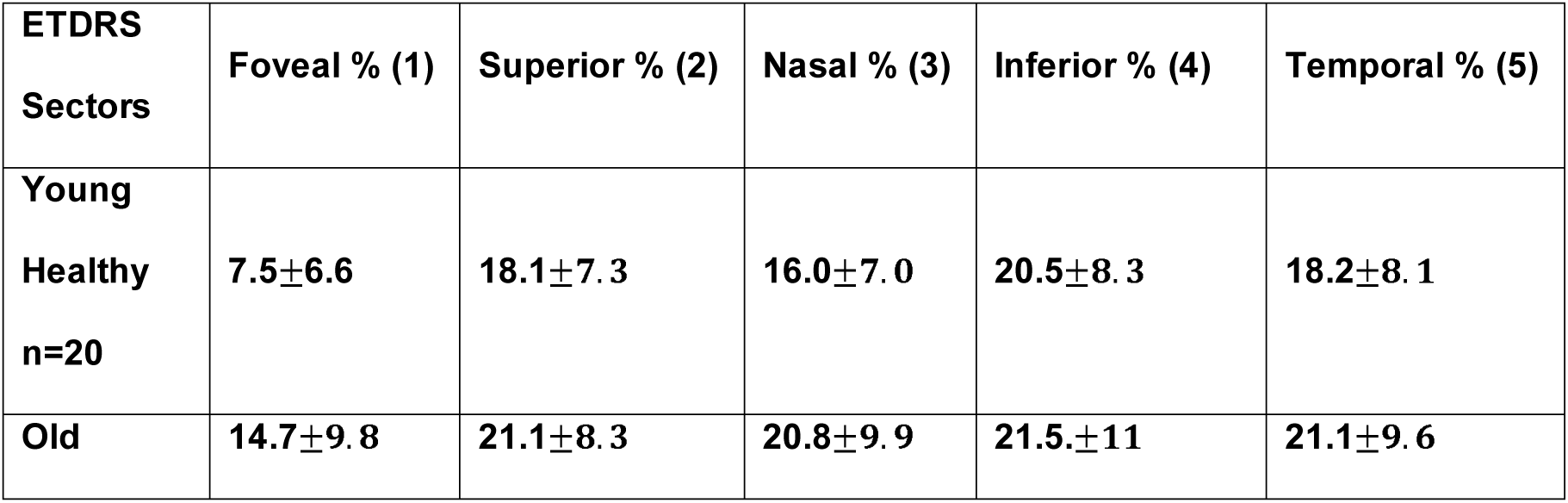
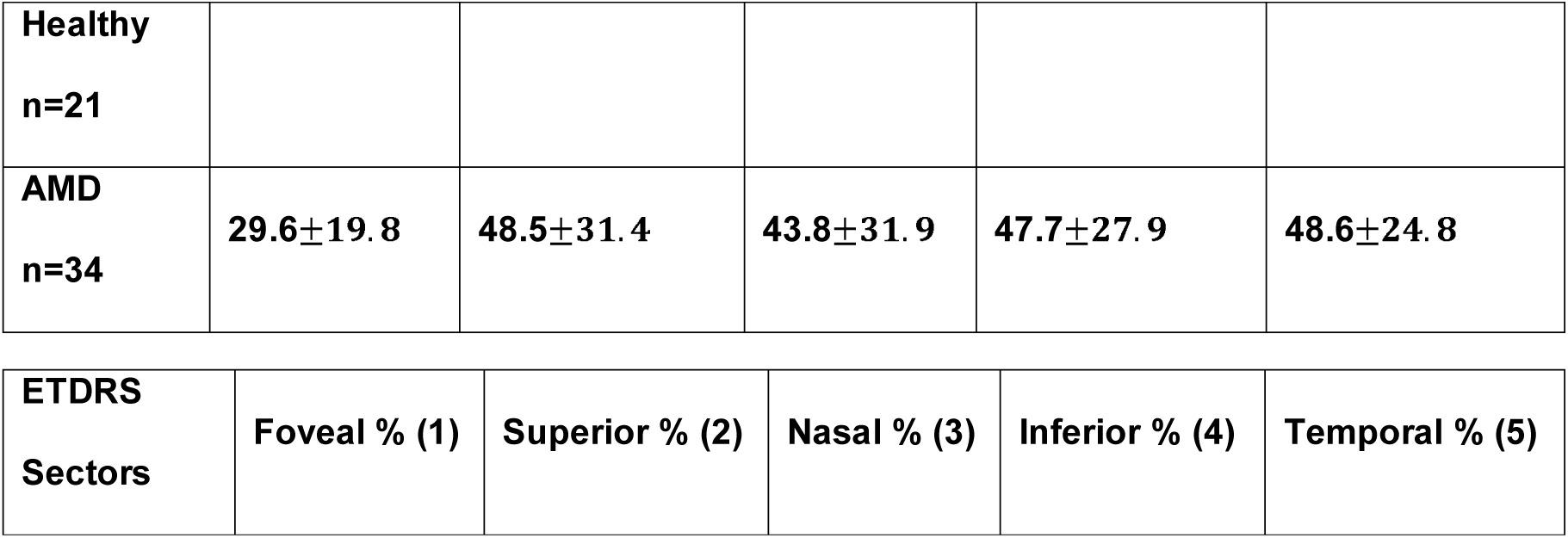
Mean Vessel Density from full retinal thickness per ETDRS Sector, for each group.

**Vessel density measurements: Full retinal thickness.**
| **ETDRS Sectors** | **Foveal % (1)** | **Superior % (2)** | **Nasal % (3)** | **Inferior % (4)** | **Temporal % (5)** |
| --- | --- | --- | --- | --- | --- |
| **Young Healthy**<br>**n=20** | **$7.5 \pm 6.6$** | **$18.1 \pm 7.3$** | **$16.0 \pm 7.0$** | **$20.5 \pm 8.3$** | **$18.2 \pm 8.1$** |
| **Old** | **$14.7 \pm 9.8$** | **$21.1 \pm 8.3$** | **$20.8 \pm 9.9$** | **$21.5 \pm 11$** | **$21.1 \pm 9.6$** |

|  |  |  |  |  |  |
| --- | --- | --- | --- | --- | --- |
| Healthy<br>n=21 |  |  |  |  |  |
| AMD<br>n=34 | 29.6±19.8 | 48.5±31.4 | 43.8±31.9 | 47.7±27.9 | 48.6±24.8 |
| ETDRS<br>Sectors | Foveal % (1) | Superior % (2) | Nasal % (3) | Inferior % (4) | Temporal % (5) |

### Vessel density measurements: SCP Segment

Table 2 shows the mean VD of the SCP segment within each ETDRS sector in each group. Although, there was a trend towards the mean vessel density derived from the SCP in all ETDRS sectors being higher in the AMD cohort compared to the 2 healthy cohorts, no significant difference was found in the VD’s recorded in any of the 5 ETDRS sectors between the 3 cohorts.

**Table 2:**
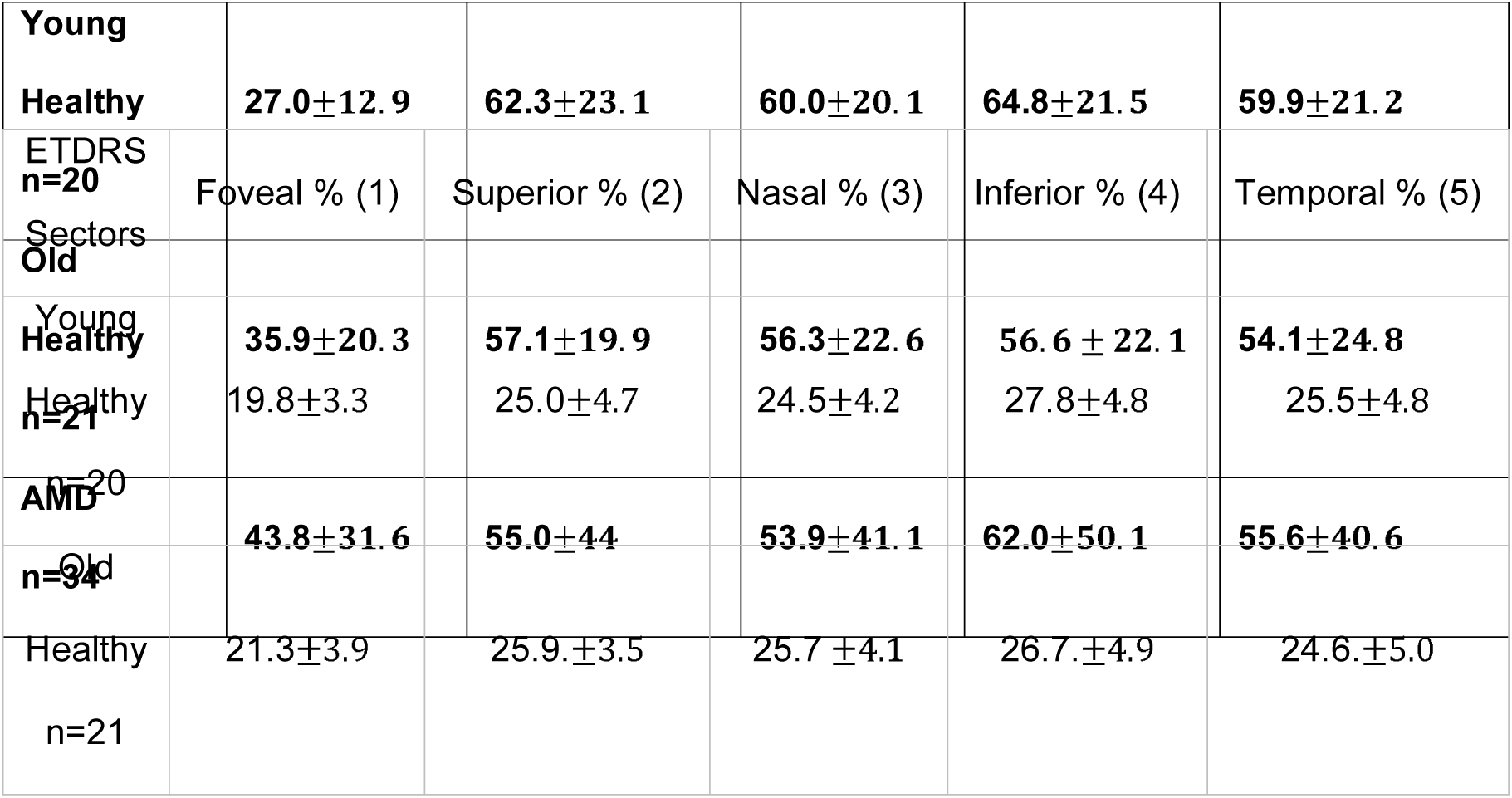
Mean Vessel Density in superficial capillary plexus (SCP) defined in sectors based on the ETDRS chart (Foveal, Superior, Nasal, Inferior, and Temporal).

### Vessel density measurements: DCP Segment

Table 3 shows the mean VD of the DCP segment within each ETDRS sector for all groups. The mean VD was found to be significantly higher (<0.01) in the DCP than the SCP in all ETDRS sectors and in all groups. Although, there was a trend towards the mean vessel density derived from the DCP in ETDRS sector 1 being higher in the AMD cohort compared to the 2 “healthy” cohorts, the difference was not statistically significant. There was a trend for the mean vessel density derived from the DCP in the remaining ETDRS sectors; 2-5, to be higher in the YH cohort compared to the other 2 cohorts, this difference was not statistically significant.

**Table 3:** Mean Vessel Density in deep capillary plexus (DCP) defined in sectors based on the ETDRS chart (Foveal, Superior, Nasal, Inferior, and Temporal).

|  |  |  |  |  |  |
| --- | --- | --- | --- | --- | --- |
| AMD | 25.4 ±7.9 | 30.4±10.5 | 30.7±11.1 | 32.5±11.0 | 32.0±8.5 |
| ETDRS | Foveal ratio | Superior ratio | Nasal ratio | Inferior ratio | Temporal ratio |

### Vessel density measurements: Choriocapillaris Segment

Table 4 shows the mean VD of the Choriocapillaris segment within each ETDRS sector for all groups. In contrast to the measurements recorded in the retinal segments, statistically significantly lower mean VD’s were recorded in the choriocapillaris segment layer in the AMD group across all ETDRS sectors compared to the two healthy groups. There was no significant difference in the VD’s measured in each of the ETDRS sectors between YH and the OH groups.

**Table 4:** Mean Vessel Density in the choriocapillaris, based on the ETDRS chart (Foveal, Superior, Nasal, Inferior, and Temporal)

| S<br>Sector<br>s | (1) | (2) | (3) | (4) | (5) |
| --- | --- | --- | --- | --- | --- |
| Young<br>Health<br>y<br>n=20 | 97 ± 10.5 | 86.2 ± 10.7 | 90.8 ± 11 | 103.8 ± 9.8 | 103 ± 11.3 |
| Old<br>Health<br>y<br>n=21 | 97 ± 4.5 | 85.5 ± 8.5 | 83.9 ± 9.5 | 86.3 ± 10.5 | 80.1 ± 12.8 |
| AMD<br>n=34 | 71.4 ± 24.3 | 68.1 ± 23.8 | 69.7 ± 24.4 | 68.4 ± 22.7 | 73.1 ± 19.4 |

## Discussion

Although a number of changes within the ageing macular have previously been reported, a reliable, non-invasive biomarker of both “healthy” macular ageing and disease risk remains elusive. Retinal thickness, both at the fovea and extra-foveal regions, has previously been shown to differ significantly between individuals with early AMD and age-matched controls with the retina being thinner in individuals with disease ^16^.The results of our study also revealed that there was a trend for the overall retinal thickness to thin with age but this difference was not statistically significant between cohorts. It is also recognised that the choroid tends to thin with age; with the nasal area being thinnest, and the subfoveal region being thickest^17^. In addition to thinning with “normal ageing,” Choroidal thinning has also been demonstrated in individuals with early AMD compared to age matched controls^16, 18-20^ however, the significance of this observation is controversial as both the extent and the pattern of thinning is significant in some^20^ but not all studies ^16,18,19^. The histological evidence regarding choroidal thickness is also contradictory with some studies showing reduction in late AMD^21^ and others showing no change^19^; moreover, the changes in the vasculature of the choriocapillaris in early AMD that have been reported from histological studies ^19, 21^ appear to be independent of choroidal thickness.

Similarly inconclusive observations have been reported with respect to the width of the Foveal avascular zone with age. Whilst some studies report no difference with age^22, 23^ others have found that the FAZ enlarges with increasing age ^24, 25^. Where reported, the FA at the level of the SCP has been found to be significantly smaller in >60 year olds compared to younger study groups^14^, but the significant of this difference is unknown and furthermore no difference was observed when the FAZ at the level of the DCP was analysed^14^. Recently, a study studying the FAZ in patients with intermediate non-exudative AMD found no significant difference in the width of the FAZ between patients with non-exudative AMD and healthy controls^26^. We likewise found there to be no statically significant difference in the diameter of the FAZ between any of the 3 cohorts studied.

Where objective measures of macular anatomy have failed, OCTA may provide an opportunity to develop a reliable, non-invasive vascular biomarker of the ageing macular. The proprietary OCTA software now has a vessel density function which, in brief uses the proportion of bright pixels to the proportion of dark pixels to derive a measure of vascular density. In the first instance we reviewed the mean vessel density percentage throughout the ***full*** retinal thickness, for each ETDRS sector. We then used the ***segmentation*** function of the enface OCTA to explore the relationship of the vessel densities within each of the ETDRS sectors 1-5 in each of the 3 vascular zones of the central macular; (SCP, DCP, Choriocapillaris) across the 3 cohorts; YH, OH and AMD.

As one might expect the VD’s derived from the relatively avascular ETDRS sector 1 were, in the ***full*** thickness maps, significantly reduced compared to the VD’s derived from ETDRS sectors 2-5 in the YH, OH, and AMD cohorts. However, and curiously, the VD recorded in the AMD cohort was significantly higher than in the two “Healthy” cohorts, and whilst there was a trend for the VD’s derived from the YH cohort to be reduced compared to the OH cohort, this difference was not significant. Aside the marked difference between the AMD cohort and the other two “healthy” cohorts, no other clinically relevant or statistically significant patterns were noted in the VD’s derived from the full thickness retinal maps.

Overall, and when the retinal vascular complexes were analysed as separate segments, the highest vessel densities were found in sectors 2 and 4; the superior and inferior macula. Again the VDs derived from ETDRS sector 1 was greater in the AMD cohort compared to the two “Healthy” cohorts. Whilst there was a trend for the VD’s derived from the OH cohort to be lower compared to the YH cohort, this difference was not significant. The spatial variation in VD’s that we observed; a higher mean VD in the inferior and superior EDTRS sectors and a lower mean VD in the temporal and nasal sectors, in both the deep and superficial capillary plexi, is consistent with published data from previous studies.^14, 22, 23^ Like others^14, 27^, we also observed that in all ETDRS sectors and in all 3 study groups, the mean VD derived from the DCP was significantly higher than that derived from the SCP.

The association between age and vessel density remains unclear, with reports of statistically significant lower mean vessel density in SCP, DCP and full thickness of retina in each ETDRS sector in patients >60 years old compared to younger cohorts;^14^, however, others who have studied this have found no such correlation^22^. On the whole, our results do not further our understanding of VD changes with age, being similarly inconclusive. On the basis of these data, one has to conclude that without further work, VD’s derived from the retinal segments are not a reliable biomarker for “healthy” ageing.

One consistent feature of the VD’s derived from the retinal segments in the present study was the finding that the VD’s recorded from ETDRS sector 1, and indeed in many other ETDRS sectors in the AMD cohort, were as a group consistently higher than those obtained from the “healthy” cohorts. One possible explanation for this surprising result is the technical challenges of obtaining high quality OCT-A scans from individuals with even modest visual impairment as accurate central fixation during the OCT-A capture can be challenging. Many of the patients in our AMD cohort struggled to maintain fixation during the image acquisition stage, despite the majority recording a Snellen acuity of 6/12.. Increased acquisition time and the resultant compromise in image quality means there is a higher likelihood of projection and banding artefacts.^28^ Motion artefacts can also lead to the duplication of inner retinal vascular structures ^29^.As vessel density is derived from the percentage of the area occupied by bright pixels in a segmented area, errors such as vessel duplication and banding artefacts could be incorrectly interpreted during computational analysis as increased vessel density within the image. Such an error would be magnified in ETDRS sector 1 where the effects of these banding and motion artefacts would be more pronounced in what should be avascular tissue.

Although the proprietary OCTA software to assess VD is designed to measure the inner retinal plexi, a vascular bed which has a very different architecture to the choriocapillaris^30^ we were curious to know whether the principle of VD analysis could be used to assess the vascularity of the choriocapillaris. We therefore extracted the raw data, and then used a manual analysis method previously published in the literature^15^, to assess the “Vascular density” of the choriocapillaris segment. Even allowing for the influence of banding and motion artefacts, issues which should increase VD measurements, we observed a significant reduction in the VD in both the AMD and OH cohorts compared to the YH cohort with the reduction in VD being most marked in the AMD cohort (Table 4). This contrasts sharply with the data derived from the retinal plexi which broadly showed no difference between groups. Although interesting, one has to interpret these findings with caution. It is possible that the observed reduction in VD is simply a result of increased projection artefact and as VD has, to the best of our knowledge, not previously been used to assess the choriocapillaris, it is not possible to corroborate these findings. However, a global reduction in choriocapillaris density, associated with drusen on OCT-A in dry AMD has been demonstrated previously.^6^ It is also recognised that the inner choroid and the choriocapillaris disproportionally thin with age and disease^31^. It is feasible that a thinning choriocapillaris would be less vascular and other groups have hypothesised that choroidal perfusion changes on the OCT-A, may predict disease progression^6^. Debate about whether this is related to actual flow impairment^6^ or projection artefact “masking” the choriocapilaris^29, 32^ due to large drusen or pigment epithelial detachments attenuating the OCT-A signal is still ongoing. Without further corroborative work, the importance of our finding that there is a progressive reduction in choriocapillaris VD in age and disease is difficult to assess. Furthermore, because our old healthy group and AMD cohorts are not aged matched, it is not possible to comment on whether any observed reduction is simply an ageing phenomenon or a marker of disease. Nevertheless, if reproducible, such a measure has the potential to provide a non-invasive biomarker of both “healthy” macular ageing and disease risk. Further limitations of this study include a small sample size, which might reduce the generalisability of our findings and the interpretation of the statistical significance of our data. Furthermore, the Kolmogorov and Smirnov statistical test assumes the Gaussian distribution of the tested data.

## Conclusion

OCT-A mean retinal vessel density results for normal subjects are in line with the existing literature. Challenges of image acquisition and the significant influence of image artefact on post-acquisition analysis of patients with mild visual loss due to intermediate age related macular degeneration further limits the value of VD measurements in this patient group. Traditional measures of vessel density, derived from the retinal plexi are therefore unreliable biomarkers for both “healthy” ageing and quantifying risk of developing AMD. Our non-proprietary analysis of the vascular density of the choriocapillaris revealed a significant drop off of VD with age and disease but further work is required to corroborate this finding. If repeatable choriocapillaris VD may provide a non-invasive biomarker of healthy ageing and disease.

## Data Availability

Data can be made available by reasonable request from authors

## Acknowledgments

The authors wish to acknowledge the assistance by Mr Diego Sonderegger, from Device Technologies®.

## Notes

### Competing Interest Statement

The authors have declared no competing interest.

### Funding Statement

no external funding was received

### Author Declarations

All relevant ethical guidelines have been followed and any necessary IRB and/or ethics committee approvals have been obtained.

Any clinical trials involved have been registered with an ICMJE-approved registry such as ClinicalTrials.gov and the trial ID is included in the manuscript.

